# Latent Class Analysis of Adult Trauma Patients Identifies Distinct Care Pathways: A Retrospective Cohort Study

**DOI:** 10.1101/2025.11.20.25340650

**Authors:** Liam Watts, Fiona Boland, Louise Brent, Pamela Hickey, Siobhan Masterson, Rory Quinn, Olga Brych, Jan Sorensen, Brid Moran, Bairbre O’Sullivan, David Willis, David Hennelly, Conor Deasy, Frank Doyle

**Affiliations:** Department of Health Psychology, School of Population Health, RCSI University of Medicine and Health Sciences, Dublin, Ireland; National Office of Clinical Audit, Dublin, Ireland; National Ambulance Service, Health Service Executive, Ireland; Spinal Injuries Ireland, Dublin, Ireland; Cork University Hospital & University College Cork, Cork, Ireland

**Keywords:** Trauma, Major trauma, Latent class analysis, Patient subgroups, Clinical trajectories, Care pathways

## Abstract

**Background:** Major trauma is a highly heterogeneous clinical condition, posing significant challenges for accurate diagnosis, timely transfer, and effective treatment. Identification of meaningful subgroups using Latent Class Analysis (LCA) can inform clinicians and policymakers, but this approach has seldom been applied to diverse trauma cohorts. We therefore aim to identify clinically meaningful subgroups of trauma patients and explore predictors of group membership.

**Methodology:** We merged data for n=4,403 patients from ambulance service electronic patient care reports with a national major trauma audit and applied LCA to identify subgroups. We then used multinomial regression to examine associations between class membership and prehospital physiological parameters.

**Results:** Using LCA, we identified five distinct patient classes by integrating demographics, care pathways, and outcomes of ranging severity: (1) Severe Trauma - critical care, (2) Moderate limb trauma - surgical management, (3) Minor-moderate chest, limb and spinal trauma - non-operative, (4) Head trauma - conservative management and (5) Complex, chest and head trauma. The five-class solution showed the best separation based on fit statistics and clinical interpretation. Analysis of prehospital physiological data showed that the Glasgow Coma Score was significantly lower in (1) and (4) with means of 13.0 and 13.2 respectively.

**Conclusions:** This first application of LCA to major trauma patients demonstrates its potential for identifying distinct patient cohorts within a heterogeneous population. Recognising these trajectories enables targeted evaluation of prehospital and in-hospital factors associated with outcomes.

## Introduction

Physical trauma remains a major cause of global ill-health, accounting for a high burden of disability-adjusted life years (DALYs) [1]. The most recent Global Burden of Disease study confirms that injuries such as road traffic accidents, falls and violence remain major contributors to mortality and reduced life expectancy [2]. In Europe, trauma continues to have a profound societal and healthcare impacts, with high costs and resource demands [3]. The epidemiological profile of severe trauma in Europe is shifting toward older populations and low-energy mechanisms, with low falls emerging as the predominant mechanism [3,4]. This shift has introduced greater clinical heterogeneity as older patients often present with different injury patterns, comorbidities and care requirements. Ireland demonstrates a similar trend, with patients aged 65 and older now representing approximately half of major trauma admissions [5].

The Injury Severity Score (ISS) [6] is the most widely used measure of trauma severity, derived from the Abbreviated Injury Scale (AIS) [7] using the three most severely injured body regions with major trauma being internationally defined as an ISS>15. However, as a single composite measure, the ISS cannot fully reflect the heterogeneity and complexity of traumatic injuries. Furthermore, its calculation requires a complete hospital-based assessment, limiting its applicability in the prehospital setting, and analysis using the ISS is usually performed retrospectively. Patients with the same ISS may follow very different clinical pathways. For example, a high-energy road traffic collision and a low-energy fall can produce similar ISS values yet lead to vastly different treatments and resource needs. Additionally, the absence of a standardised definition for “multiple trauma” contributes to variability across studies, complicating comparisons and interpretation of outcomes [8].

The journey of trauma patients through healthcare systems is a complex, multi-stage process encompassing prehospital assessment, treatment and transport, emergency department stabilisation, in-hospital management, and post-hospital discharge. Qualitative studies have shown that patients often experience this process as overwhelming and variable, influenced by factors such as prehospital interventions, communication, staff coordination and discharge planning [9-11]. Variability in patient experiences reflects differences in injury severity, mechanism, and care pathways owing to the heterogeneous nature of trauma. To overcome these limitations, quantitative clustering-based approaches offers a potential means to identify clinically meaningful trauma subgroups and patient trajectories, providing a more nuanced understanding of injury heterogeneity, thereby complementing traditional severity scores and potentially providing insights for future management.

While LCA has been applied in specific cohorts of trauma injuries, no study to date has considered heterogeneous presentations to explore typical presentations and patient journeys. Unlike traditional variable-centred methods such as regression models, which examine associations between individual predictors and outcomes, clustering-based approaches adopt a person-centred perspective to uncover naturally occurring subgroups within diverse clinical data. For example, a recent systematic review [12] of clustering-based studies in traumatic brain injury (TBI), one of the most extensively studied trauma subfields, found that few papers used LCA whereas more than half employed hierarchical clustering, which deterministically assigns individuals to groups based on distance metrics. Such methods are limited by their reliance on subjective judgement when selecting the number of clusters and by their poor suitability for categorical data. In contrast, latent class analysis is a model-based probabilistic technique that enables reproducible class assignment and appropriately accommodates categorical variables, making it a robust and transparent alternative for analysing complex clinical populations. It also provides statistical measures of model fit and classification quality while allowing clinical expertise to inform decisions regarding the optimal number of classes.

LCA has been increasingly used to characterise heterogeneity in complex in-hospital populations, including patients with sepsis [13], cancer [14], complex patient profiles [15], and multimorbidity [16]. Across these settings, it has demonstrated the capacity to identify clinically meaningful subgroups and care patterns, informing tailored interventions and providing insights that extend beyond traditional variable-centred analyses. Despite this, applications of LCA within general trauma populations remain limited. Existing studies have largely focused on specific injury types or specific patient cohorts, particularly in TBI [17-19], which is a leading cause of morbidity and mortality worldwide [20]. In addition, LCA has been used to explore the overall intensity of trauma care delivery [21] suggesting potential for broader application to heterogeneous in-hospital trauma populations. However, to our knowledge, LCA has not yet been applied to in-hospital major trauma patients to inform the patient journey. Therefore, the aim of the present study was to apply LCA to in-hospital trauma patients in Ireland to identify typical patient subgroups and associated care pathways.

## Methodology

### Study Design and Setting

We conducted a retrospective cohort study, using linked data from the Major Trauma Audit (MTA) [22] and the National Ambulance Service (NAS) in Ireland, covering patients admitted between the 1^st^ of January 2020 and the 31^st^ of December 2022. The MTA is a national clinical audit that provides standardised clinical data on trauma patients who fulfil criteria based on ISS, length of stay and injury details (Appendix A). The NAS provides prehospital emergency care and transport services across Ireland and trauma patients were identified as those with a working diagnosis containing “trauma”. Descriptive statistics were used to profile the sample.

### Data sources and Variables

In-hospital data were obtained from the MTA. Variables included:

- **Demographics:** Age and sex.
- **Injury severity and treatment:** ISS, the number of inter-hospital transfers, number of operations, and admission to critical care.
- **Clinical process variables:** Most senior clinician treating the patient in the emergency department (ED) and the most severely injured body region.
- **Outcomes:** Hospital length of stay (categorised into quartiles) and discharge destination.

Prehospital data were derived from NAS records. The first physiological variables measured included: Glasgow Coma Score (GCS), systolic blood pressure (SBP), heart rate (HR), oxygen saturation (SpO₂), and respiratory rate (RR).

### Statistical Methods

#### Latent Class Analysis

Latent Class Analysis (LCA) was conducted to identify unobserved subgroups of trauma patients based on in-hospital variables present in the MTA dataset. LCA is a latent variable modelling approach in which categorical latent classes are inferred from observed indicators, allowing the identification of distinct subgroups within heterogeneous populations. Continuous variables were categorised to enhance clinical interpretability. Age was grouped into five categories (≤35, 36–55, 56–65, 66–80, and >80 years) to preserve interpretability and ensure representation across adult life stages within the trauma cohort. Length of hospital stay was divided into quartiles to reflect increasing resource use and recovery time. The Injury Severity Score (ISS) was categorised as mild (≤ 8), moderate (9-15), and severe (>15) to align with established trauma severity thresholds. Although categorisation of continuous indicators may reduce some information, it is a common and accepted approach in latent class analysis to support meaningful class interpretation [23]. This approach also ensured compatibility with the poLCA package (version 1.6.0.1) in R (version 4.4.3), which treats all variables as unordered categorical indicators.

Model fit was evaluated using the Akaike Information Criterion (AIC), Bayesian Information Criterion (BIC), and entropy, alongside class membership probabilities. For each individual, class membership probabilities represent the likelihood of belonging to each latent class given their observed data, with 1/k (where k is the number of classes) indicating maximal uncertainty and values approaching 1 indicating high certainty of class assignment [23]. The class probability, or the proportion of patients assigned to each class, was calculated to describe class sizes. The average posterior probability for each class was calculated to summarise classification confidence, with values above 0.8 considered indicative of good separation. The minimum and maximum average posterior probabilities across classes were also examined to ensure that even the least certain class was well-classified [23, 24]. Entropy, which ranges from 0 to 1, was used to assess overall classification certainty, with values greater than 0.8 commonly considered indicative of good separation, though this should be interpreted alongside other model fit indices and the substantive interpretability of the classes [23, 24]. Each latent class was required to include at least 10% of patients to ensure stability, interpretability and adequate representation of the trauma population [23]. While smaller classes could represent clinically distinct subgroups, they may not provide a sufficiently large or representative cohort for educational or translational purposes.

### Multinomial Regression

Following LCA, multinomial logistic regression was used to examine whether prehospital physiological variables were associated with LCA-derived class membership. The outcome variable was class membership, with Class 1 used as the reference category, and all predictor variables were modelled as linear predictors.

Analyses were restricted to patients with complete prehospital data, to avoid bias from missingness patterns. Descriptive statistics (mean and standard deviations) were reported for each variable, along with relative risk ratios (RRRs) and 95% confidence intervals (CIs). Regression analyses were performed in Stata version 19.5.

### Potential Bias

Potential sources of bias were considered. Prehospital data were only available for patients transported by the NAS, representing a subset of the full MTA dataset. To assess the impact of this limitation, LCA was performed on the entire MTA dataset and compared with the subset of patients transported by the NAS. Although the NAS operates throughout Dublin, the Dublin Fire Brigade (DFB) also provides ambulance services in the area; as DFB records remain paper-based, these patients were not included in the study.

### Patient and public involvement statement

This study was a retrospective analysis of linked national major trauma audit and ambulance service data. Patients and the public were not involved in the design, conduct, or reporting of this research, as it relied solely on existing clinical records.

## Results

### Participants and study size

We included all adult patients (aged ≥ 16 years) recorded in the MTA between 2020 and 2022 (n=11,354). Of these, n=4,470 patients were transported to the hospital by the NAS and successfully linked to their corresponding prehospital records. All patients were treated in designated Major Trauma Centres (MTCs) or Trauma Units (TUs), corresponding to Level I and Level II trauma centres. A STROBE flow diagram detailing the cleaning process is shown in Figure 1.

**Figure 1:**
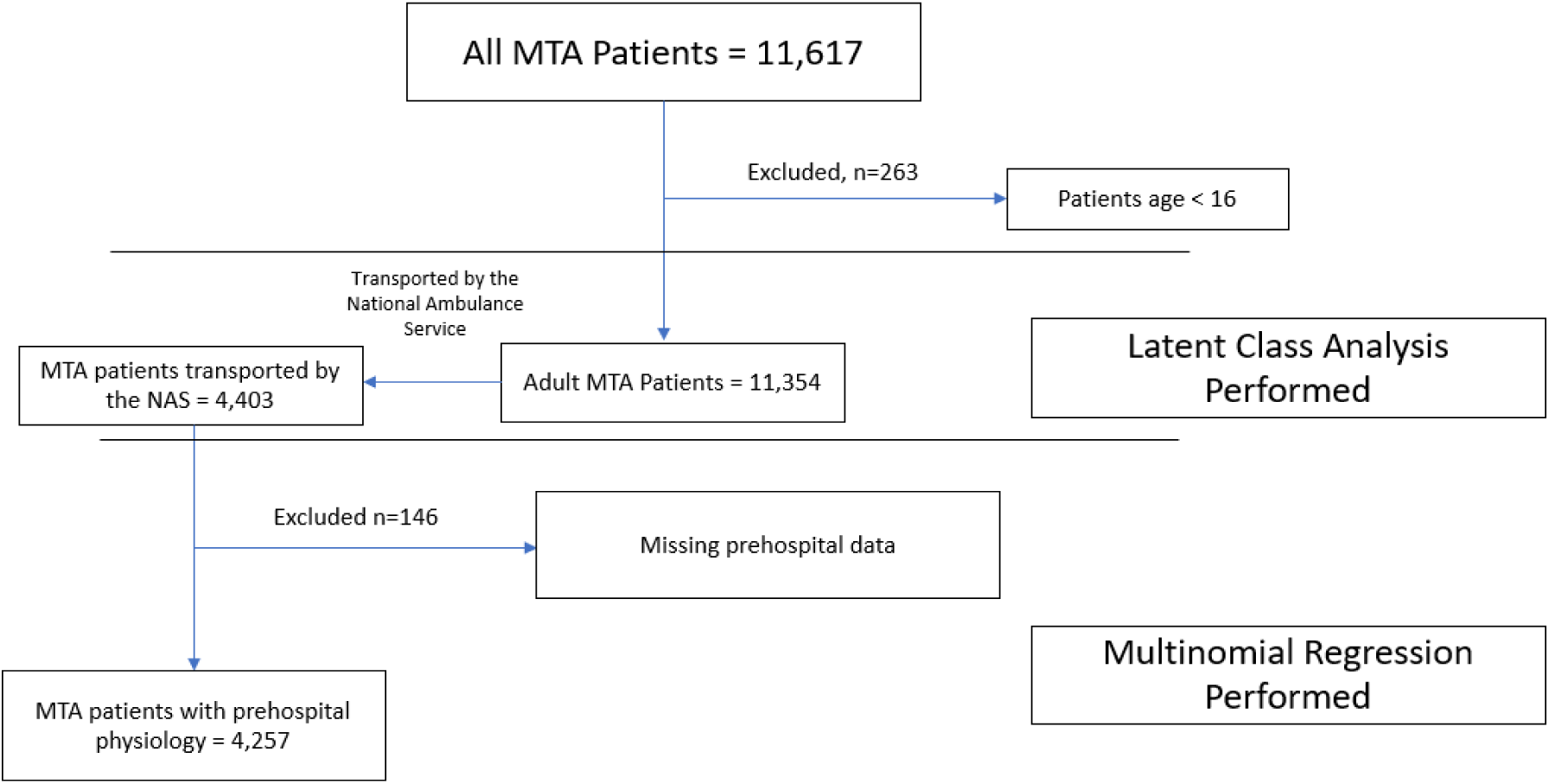
STROBE flow diagram showing patient inclusion and cohort derivation for LCA and regression analyses.

### Descriptive Data

Table 1 shows the descriptive statistics of the MTA patients transported to hospital via the NAS (n=4,403). The cohort was predominantly male with a mean age of ∼65 years. Injury severity was most commonly moderate or severe, and the majority of patients were discharged to home-based care. The distribution of the most severely injured body regions reflected a range of categories, with the “Other” category capturing injuries not otherwise specified in the primary classifications.

**Table 1:** Descriptive statistics for patients transported to hospital by the NAS.

| Total (n=4,403) | Variable |  |
| --- | --- | --- |
| **Demographics** | Age, mean(SD) | 65.3 (20.1) |
|  | Male, n(%) | 2,362 (53.7%) |
| **Hospital Variables** | ISS (mild), n(%) | 813 (18.5%) |
|  | ISS (moderate), n(%) | 2,086 (47.4%) |
|  | ISS (severe), n(%) | 1,504 (34.2%) |
| | Transfers $\geq 1$, n(%) | 534 (12.1%) |
| | Operations $\geq 1$, n(%) | 1,794 (40.1%) |
|  | Critical Care Admission, n(%) | 623 (14.2%) |
| **Most Senior ED** | Consultant, n(%) | 926 (21.0%) |
|  | Registrar, n(%) | 2,123 (48.2%) |
|  | Specialist Registrar, n(%) | 516 (11.7%) |
|  | Senior House Officer, n(%) | 838 (19.0%) |
| **Most Severely Injured**<br>**body area** | Chest, n(%) | 705 (16.0%) |
|  | Head, n(%) | 967 (22.0%) |
|  | Limbs, n(%) | 1,455 (33.1%) |
|  | Spine, n(%) | 687 (15.6%) |
|  | Other, n(%) | 589 (13.4%) |
| **Discharge Destination** | Home-based care, n(%) | 2,556 (58.1%) |
|  | Institutional care, n(%) | 854 (19.4%) |
|  | Rehabilitation, n(%) | 690 (15.7%) |
|  | Mortuary, n(%) | 303 (6.9%) |
| **Prehospital Variables** | Glasgow Coma Score, mean(SD) | 14.2 (2.9) |
|  | Systolic Blood Pressure, mean(SD) | 141.2 (28.6) |
|  | Heart Rate, mean(SD) | 84.9 (20.5) |
|  | Oxygen Saturation, mean(SD) | 94.8 (6.7) |
|  | Respiration Rate, mean(SD) | 19.1 (6.5) |

### Model Selection

Comparative model fit statistics for 2-10 classes, based on the variables presented in Table 1 (excluding prehospital physiological variables), are shown in Table 2. All models converged normally, and comparable fit statistics for the full MTA cohort are provided in Table A1 (Appendix B).

**Table 2:** Fit statistics for 2-10 latent class solutions for MTA patients with prehospital physiology.

| Class # | Entropy | Akaike<br>Information | Bayesian<br>Information | Average Class Probability |  |
| --- | --- | --- | --- | --- | --- |
|  |  | Criterion (AIC) | Criterion (BIC) | Minimum | Maximum |
| 2 | 0.75 | 95767.06 | 96131.294 | 0.927 | 0.931 |
| 3 | 0.853 | 92725.89 | 93275.431 | 0.921 | 0.957 |
| 4 | 0.868 | 90607.85 | 91342.704 | 0.894 | 0.962 |
| **5** | **0.879** | **89272.58** | **90192.749** | **0.893** | **0.961** |
| 6 | 0.869 | 88626.45 | 89731.927 | 0.843 | 0.953 |
| 7 | 0.867 | 88115.19 | 89405.974 | 0.832 | 0.94 |
| 8 | 0.868 | 87813.22 | 89289.322 | 0.844 | 0.942 |
| 9 | 0.861 | 87556.87 | 89218.278 | 0.833 | 0.928 |
| 10 | 0.864 | 87320.31 | 89167.034 | 0.82 | 0.943 |

The 5-class solution was selected as the optimal model, based on a combination of statistical fit and clinical interpretability. Entropy peaked for the 5-class solution, indicating the greatest separation among classes. The minimum average class posterior probability also exceeded that of the 6-class solution, supporting reliable individual classification. Class membership was assigned based on the highest posterior probability, and although this approach introduces some classification uncertainty, the average posterior probabilities (>0.8) indicated good separation across classes

Although AIC and BIC continued to decline with additional classes, the improvements became progressively smaller beyond five classes, indicating that additional classes provided limited gains in statistical fit. The 5-class solution provided the most clinically meaningful distinction between patient profiles. Class-specific conditional probabilities (Table 3) revealed that each class was substantively interpretable, whereas the 4-class solution offered less granularity, and the 6-class solution included classes with lower assignment certainty and overlapping clinical characteristics (Tables A3 and A4, Appendix C). Comparison of the 5-class solution in Table 3 with the class specific conditional probabilities for the entire MTA cohort (Table A2 Appendix B) revealed the same defining patient characteristics.

**Table 3:**
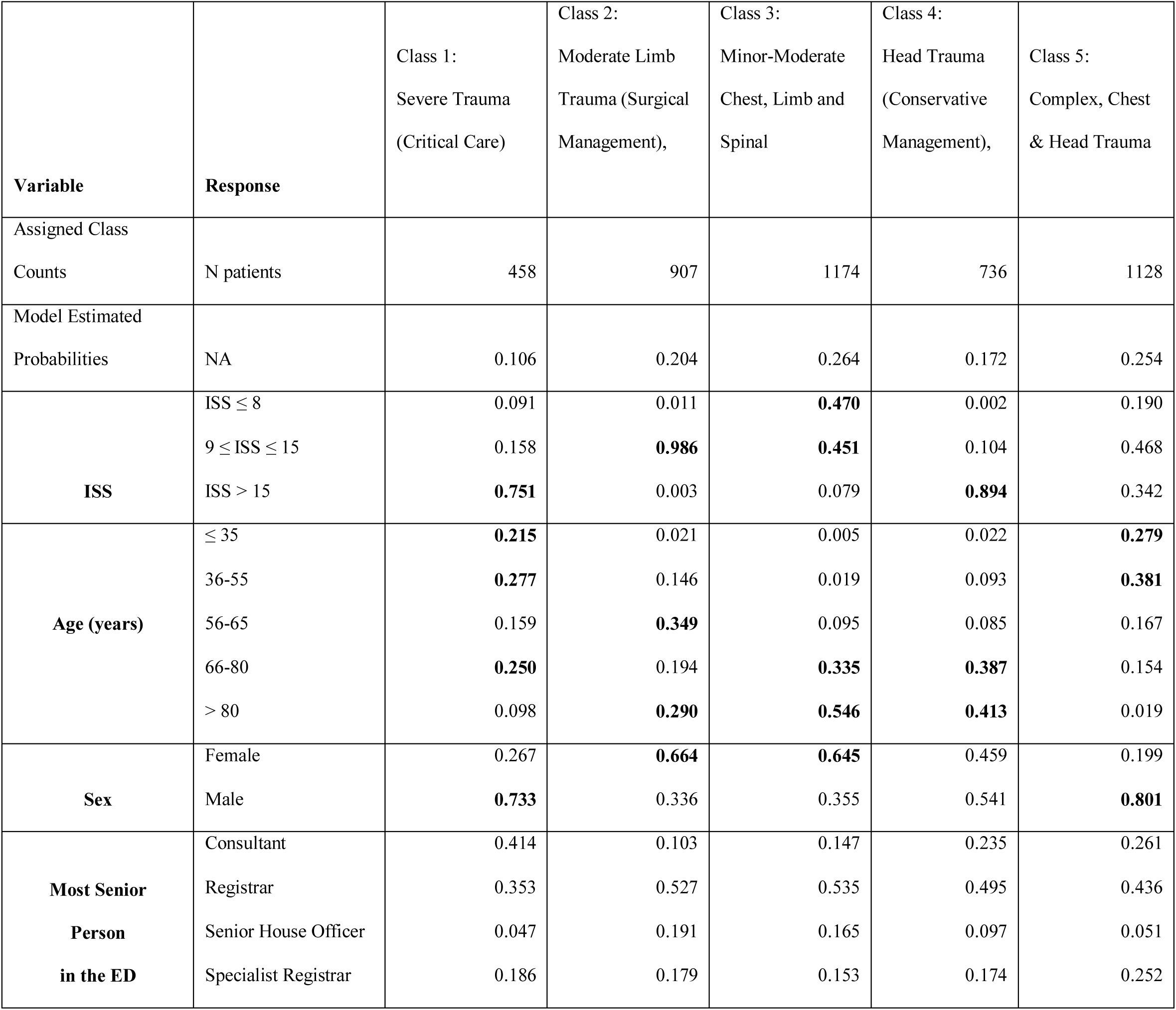
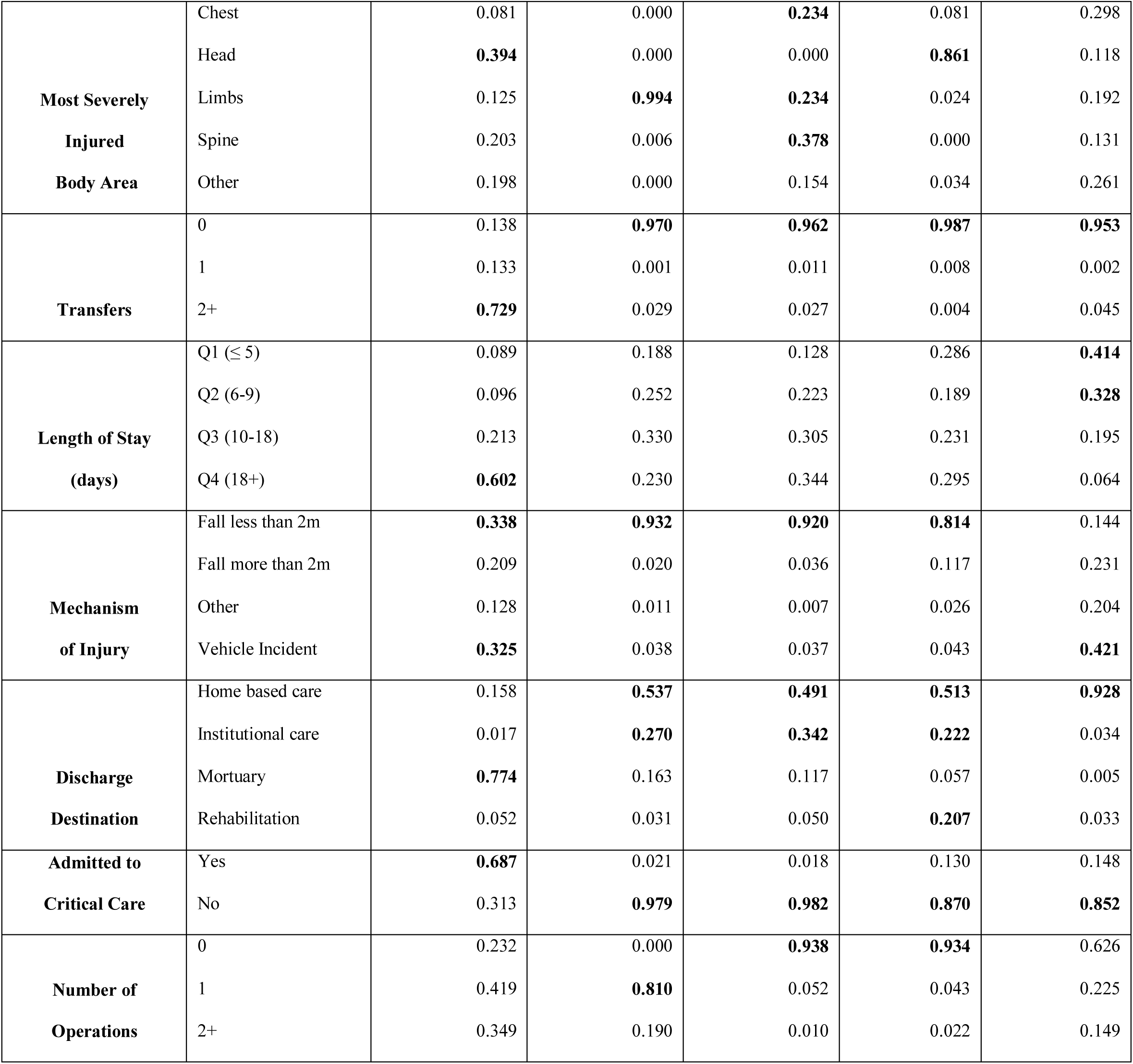
Class specific conditional probabilities for the five-class solution of MTA patients with prehospital physiology.

### Latent classes

Table 3 presents the class specific conditional probabilities for the optimal 5-class solution.

Class 1, labelled as *Severe Trauma (Critical Care),* comprised the most severely injured patients, with 75% having an Injury Severity Score (ISS) >15. This class included predominantly younger and middle-aged males, as well as some aged 66-80 years. Mechanisms of injury were mainly low falls and motor vehicle collisions. These patients were frequently transferred, often admitted to critical care, had prolonged hospital stays, and were most commonly discharged to the mortuary.

Class 2, labelled as *Moderate Limb Trauma (Surgical Management),* consisted almost exclusively of patients with limb injuries of moderate severity (ISS ranging 9-15), predominantly middle-aged and older females following low falls. Most were managed by registrars, typically underwent a single operation, and approximately half were discharged home.

Class 3 included older patients with minor to moderate injuries involving the spine, limbs, or chest, most often resulting from low falls. The majority were discharged either home or to institutional care. We labelled this class as *Minor-Moderate Chest, Limb and Spinal*.

Class 4, labelled *Head Trauma (Conservative Management),* represented older, severely injured patients with head injuries sustained predominantly from low falls. These patients demonstrated variable lengths of stay and were discharged across home-based, institutional, and rehabilitation care settings, with rehabilitation being more common in this class than other classes.

Class 5 included generally younger male patients with less severe injuries compared to Class 1. The predominant mechanism of injury was vehicle-related, with few admitted to critical care and the vast majority discharged home. This class we labelled as *Complex, Chest & Head Trauma*.

### Prehospital Variables

Table 4 presents the results of the multinomial logistic regression examining the association between prehospital physiological variables and latent class membership, using Class 1: Severe Trauma as the reference category. Across models, GCS and SpO₂ demonstrated the most consistent and statistically significant associations with class membership whereas SBP, RR, and HR showed smaller effects. GCS was significantly higher in Classes 2, 3, and 5 compared with Class 1, and not significantly different in Class 4. SBP was significantly higher in Classes 2, 3, and 4 but not in Class 5. RR was significantly lower in Classes 2-4 compared with Class 1. HR showed no significant differences across classes except for Class 5. SpO₂ was significantly higher in Class 3, with no significant differences observed for the other classes.

**Table 4:** Multinomial regression results showing associations between prehospital physiology and latent class membership with Class 1 as the reference category. Displayed are mean (SD), relative risk ratio (95% CI) and p-value.

|  |  |  |  |  |  |
| --- | --- | --- | --- | --- | --- |
| **Class** | **Reference: (1)**<br>**Severe Trauma**<br>**(Critical Care)** | **(2) Moderate**<br>**Limb Trauma**<br>**(Surgical**<br>**Management)** | **(3) Minor-**<br>**Moderate Chest,**<br>**Limb**<br>**and Spinal (Non-**<br>**Operative)** | **(4) Head**<br>**Trauma**<br>**(Conservative**<br>**Management)** | **(5) Complex, Chest**<br>**& Head Trauma**<br>**(Short Stay)** |
| **Glasgow Coma Scale**<br>**(GCS)** | 13.0 (3.5) | 14.8 (0.6)<br>RRR 1.59 (1.47,<br>1.71)<br>p <0.001 | 14.8 (2.5)<br>RRR 1.59 (1.47,<br>1.71)<br>p <0.001 | 13.2 (3.2)<br>RRR 1.00 (0.97,<br>1.04)<br>p= 0.73 | 14.5 (3.3)<br>RRR 1.26 (1.20,<br>1.32)<br>p <0.001 |
| **Systolic Blood**<br>**Pressure (SBP)** | 133.5 (28.6) | 142.5 (28.0)<br>RRR 1.01 (1.00,<br>1.02)<br>p <0.001 | 147.1 (28.1)<br>RRR 1.02 (1.01,<br>1.02)<br>p <0.001 | 148.2 (29.7)<br>RRR 1.02 (1.01,<br>1.02)<br>p <0.001 | 133.4 (26.2)<br>RRR 1.00 (0.99,<br>1.00)<br>p= 0.21 |
| **Respiration Rate (RR)** | 19.9 (9.0) | 18.4 (4.9)<br>RRR 0.96 (0.94,<br>0.98)<br>p <0.001 | 19.1 (6.9)<br>RRR 0.98 (0.97,<br>1.00)<br>p=0.01 | 18.8 (5.6)<br>RRR 0.98 (0.96,<br>0.99)<br>p=0.01 | 19.5 (6.5)<br>RRR 0.99 (0.98,<br>1.00)<br>p= 0.27 |
| **Heart Rate (HR)** | 85.0 (22.5) | 83.0 (17.8)<br>RRR 1.00 (0.99,<br>1.00)<br>p=0.25 | 84.7 (18.9)<br>RRR 1.00 (0.99,<br>1.00)<br>p= 0.75 | 83.7 (20.7)<br>RRR 0.99 (0.99,<br>1.00)<br>p=0.08 | 87.2 (21.8)<br>RRR 1.01 (1.00,<br>1.01)<br>p= 0.017 |
| **Oxygen Saturation**<br>**(SpO<sub>2</sub>)** | 94.2 (8.8) | 95.7 (5.4)<br>RRR 1.01 (0.99,<br>1.04)<br>p=0.28 | 94.2 (7.1)<br>RRR 0.97 (0.95,<br>0.99)<br>p <0.001 | 94.8 (4.8)<br>RRR 1.00 (0.98,<br>1.01)<br>p=0.82 | 95.2 (7.1)<br>RRR 1.01 (0.99,<br>1.03)<br>p= 0.56 |

### LCA Descriptions

Based on the latent class structure presented in Table 3 and the corresponding prehospital physiological characteristics shown in Table 4, Table 5 provides a summary of each class with descriptive labels in addition to clinical and demographic features. These summaries integrate injury severity, mechanism, and care pathway characteristics to provide a clear overview of the heterogeneity within the MTA trauma population.

**Table 5:** Descriptive summary of each latent class. Including key clinical, demographic and care pathway characteristics.

| Class | Label | Prehospital physiology | Patient Profile and Injury characteristics | Care Pathway |
| --- | --- | --- | --- | --- |
| 1 | **Severe Trauma - Critical Care** | Low GCS, normal to elevated vitals, slightly low SpO<sub>2</sub> | Predominantly male (73%) patients with severe ISS | Multiple transfers, long hospital stay, admitted to critical care. High mortality (77%) discharged to a mortuary). |
| 2 | **Moderate Limb Trauma - Surgical Management** | Near-normal GCS, slightly elevated SBP | Middle-aged and older female (66%) with moderate ISS from limb injuries due to low-falls. | No transfers, 81% of patients had 1 operation. Discharged mainly to Home (54%) or institutional care (27%) |
| 3 | **Minor-Moderate Chest, Limb and Spinal Trauma - non-Operative** | Near-normal GCS, slightly elevated SBP, slightly lower SpO<sub>2</sub> | Older female (65%) patients with mild to moderate ISS from chest, limb and spine injuries due to low-falls. | No transfers, not admitted to critical care. Discharged to Home (49%) or institutions (34%) |
| 4 | **Head Trauma - Conservative** | Low GCS, slightly elevated SBP | Older patients with severe ISS from head injuries due to low-falls. | No transfers, not admitted to critical care. Discharged to Home (51%), institutions (22%) or rehabilitation (21%) |
| 5 | **Complex, Chest and Head Trauma - Short Stay** | Near-normal GCS, other vitals similar to the severe trauma class | Younger, predominantly male (80%) with various ISS from chest and other injuries. MOI consisted of vehicle incidents (42%), falls more than 2m (23%) and other injuries (20%). | No transfers, not admitted to critical care. Short stay; 37% requiring an operation. Discharged Home (93%) |

## Discussion

This is the first study to apply LCA to a heterogeneous group of in-hospital major trauma patients. Our findings are novel, identifying five distinct patient pathways that explore prehospital vital signs through to in-hospital management and outcomes. The classes were consistent across both the full MTA cohort and the subset with linked prehospital data, highlighting the reproducible patterns of patients in each cohort. Interestingly, male patients were overrepresented in the most severe and least severe classes. Prehospital variables were also differentially predictive of different groups, providing further evidence of discrimination. The profiles of our five-class solution among patients with linked prehospital data align closely with the class-specific conditional probabilities for the full MTA cohort, as shown in Table A2 (Appendix B). The latent classes identified are comparable to cohorts described in previous literature.

The *Severe Trauma (Critical Care)* class is predominantly younger and male, with high ISS and frequent critical care admission, resulting in 77% of patients discharged to a mortuary. This class is comparable to the group requiring high intensity care in [21] where they are predominantly severely injured males discharged to a hospice (44%) or a skilled nursing facility (37%). Compared with recent Irish critical care trauma cohorts [25], our severe trauma class shows comparable ISS and TBI frequency.

The *Moderate Limb Trauma (Surgical Management)* class included predominantly female patients with injuries sustained from low falls. All patients underwent surgery with 81% of patients requiring only a single operation, aligning with previous studies reporting high limb salvage with single-stage procedures [26]. Single-stage surgery is recommended to shorten hospital stay and aid recovery in sustained limb injuries [27]. Our analysis reports prolonged rehabilitation with 46% of patients discharged to further care and 23% had a hospital stay of more than 18 days [28].

The *Minor-Moderate Chest, Limb and Spinal (Non-Operative)* class represented older, predominantly female patients with injuries from low falls, managed conservatively. Evidence supports non-operative management in this population, with low complication rates and good recovery [29-31]. The 46% discharged to further care highlights the ongoing need for post-acute rehabilitation.

*Head Trauma (Conservative Management)* comprised older patients with injuries sustained from low falls, managed without critical care. This mirrors trends in low-energy fall TBI admissions in older adults [32]. Current guidance supports selective non-operative care, as survival outcomes are comparable to surgical management despite high dependency and poorer well-being [33-35]. In our results, TBI spanned both Classes 1 and 4, emphasising its role across the severity spectrum, with affected patients characterised by a low GCS in the prehospital setting.

Class 5, *Complex, Chest & Head Trauma (Short Stay)* included younger male patients with moderate injuries, predominantly managed non-operatively. These patients typically experienced short hospital stays, minimal ICU utilisation, and were most often discharged to home-based care. Comparable studies [36, 37] report similar ISS values, low mortality rates, and conservative management approaches.

In our cohort, younger males involved in road traffic accidents (RTAs) were distributed between Class 1 (33%) and Class 5 (42%), illustrating a continuum of injury severity within this demographic, consistent with previous findings that younger males account for the majority of RTA-related trauma and rehabilitation admissions in Ireland [38]. Overall, these findings reflect the heterogeneity of trauma presentations across age, sex, and trauma outcomes, revealing distinct care pathways that may inform future trauma system design and targeted intervention strategies

### Strengths and Limitations

This study has several strengths. We were able to identify distinct classes of trauma patients, based on heterogeneous data. By comparing the entire MTA cohort to those transported by the NAS, we found that the key class distinctions between patient groups were consistent, demonstrating that the subset of MTA patients included in our analysis was representative of the larger population. We also acknowledge the study limitations. Since LCA generates classes based on unordered variables, continuous quantities such as ISS, age and length of hospital stay were not recognised as sequential as the package used for LCA, poLCA, treats all variables as unordered. While a five-class solution was the most clinically interpretable and had strong fit statistics, the AIC and BIC did not find a global minimum between 2-10 classes, likely reflecting the underlying heterogeneity of trauma. Additionally, data from the Dublin area for patients transported by the Dublin Fire Brigade were unavailable due to the use of a paper-record system [39] which may introduce geographic bias.

## Conclusion

This study represents the first application of latent class analysis to a heterogeneous trauma population within a trauma network, identifying five distinct patient classes with unique clinical, demographic, and care pathway characteristics. The latent classes showed effective stratification of trauma patients, revealing patterns across both prehospital and in-hospital variables. These findings provide a framework for evaluating trauma care across a range of injuries and indicators and support the design of targeted interventions and optimised care pathways tailored to each patient subgroup.

## Availability of supporting data

The data used in this study are not publicly available due to their sensitive nature and privacy concerns. The data was provided by the Major Trauma Audit and the National Ambulance Service in Ireland.

## Acknowledgements

The authors would like to thank all audit coordinators and everyone in the National Ambulance Service for their continued work in collecting and preparing data, the Health Service Executive (HSE) and Richard Murray for his Patient and Public Involvement.

## Appendix A Major Trauma Audit Inclusion Criteria

The inclusion criteria for the MTA audit can be found in the MTA report [40] with the length of stay criteria:

### Direct Admissions

- Trauma admissions whose length of stay is 3 days or more
- Trauma patients admitted to a High Dependency Area regardless of length of stay
- Deaths of trauma patients occurring in the hospital including the Emergency Department (even if the cause of death is medical)
- Trauma patients transferred to other hospital for specialist care or for an ICU/HDU bed.

### Patients Transferred in

- Trauma patients transferred into your hospital for specialist care or ICU/HDU bed whose combined hospital stay at both sites is 3 days or more
- Trauma admissions to an ICU/HDU area regardless of length of stay
- Trauma patients who die from their injuries (even if the cause of death is medical)

## Appendix B LCA fit statistics for the entire MTA dataset

This appendix presents the results of latent class analysis (LCA) applied to the entire Major Trauma Audit (MTA) cohort, including patients without prehospital physiological data. While the main analysis focused on patients with complete prehospital physiology, Table A1 shows the fit statistics and Table A2 shows the class specific conditional probabilities for the five-class solution.

**Table A1:** LCA fit statistics for the entire MTA dataset (n=11,354)

| Class # | Entropy | Akaike<br>Information<br>Criterion (AIC) | Bayesian<br>Information<br>Criterion (BIC) | Average Class Probability |  |
| --- | --- | --- | --- | --- | --- |
|  |  |  |  | Minimum | Maximum |
| 2 | 0.741 | 252511.751 | 252915.304 | 0.924 | 0.929 |
| 3 | 0.846 | 245996.762 | 246605.76 | 0.927 | 0.945 |
| 4 | 0.859 | 240073.649 | 240888.092 | 0.898 | 0.946 |
| **5** | **0.862** | **236896.261** | **237916.149** | **0.886** | **0.947** |
| 6 | 0.851 | 235172.584 | 236397.917 | 0.836 | 0.948 |
| 7 | 0.862 | 233598.629 | 235029.407 | 0.833 | 0.948 |
| 8 | 0.866 | 232338.887 | 233975.111 | 0.832 | 0.947 |
| 9 | 0.86 | 231339.752 | 233181.42 | 0.817 | 0.914 |
| 10 | 0.865 | 230479.053 | 232526.167 | 0.815 | 0.915 |

**Table A2:**
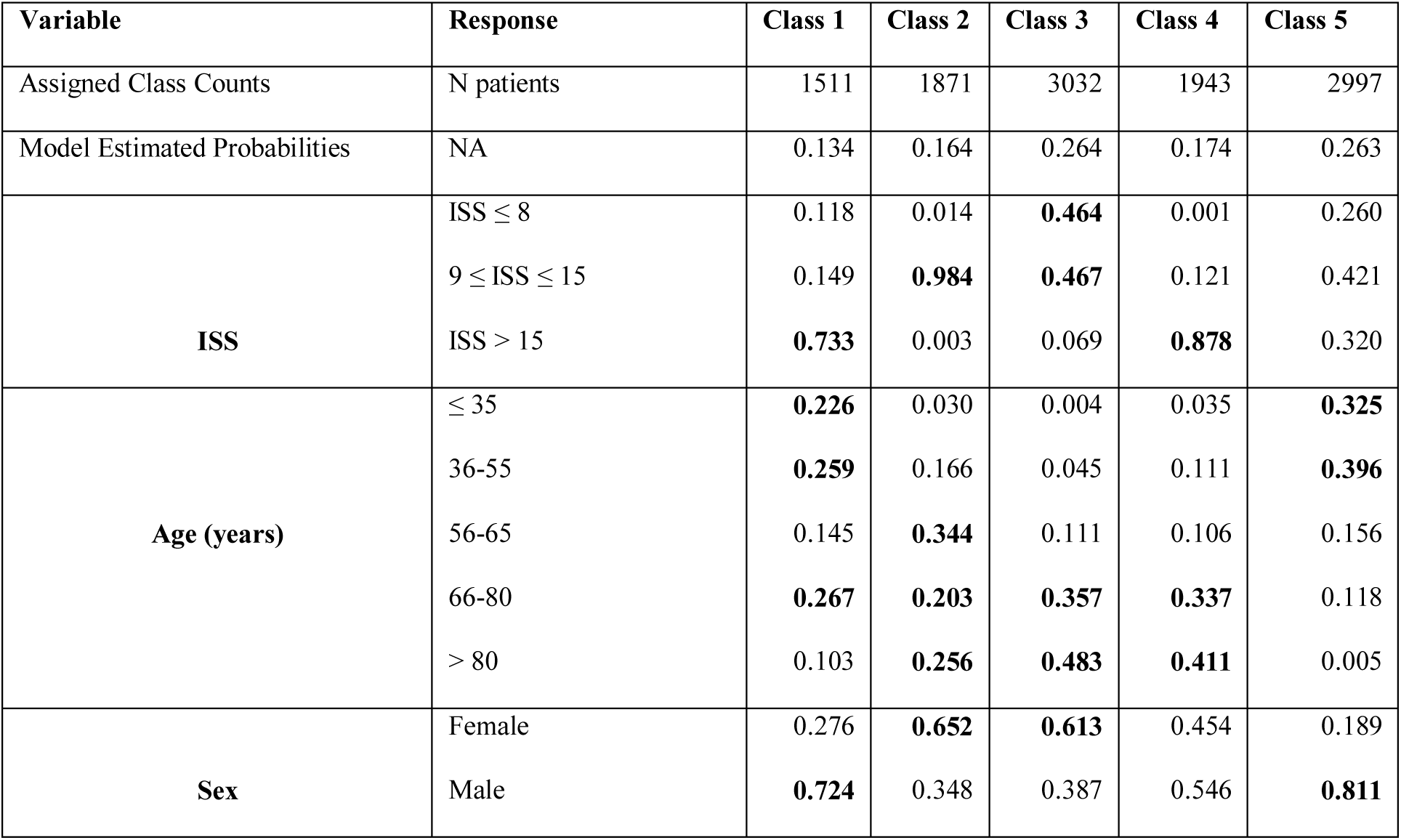
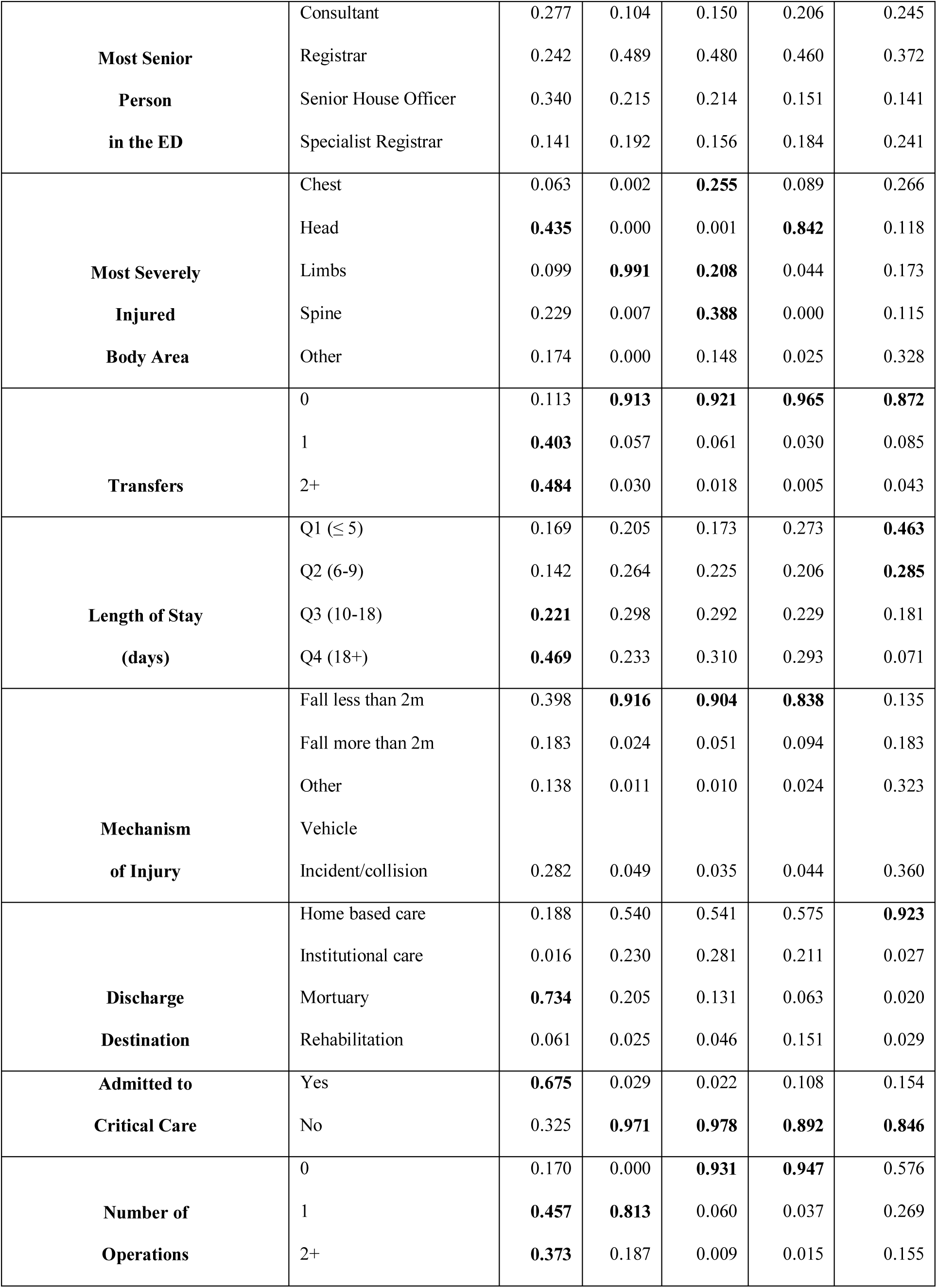
Class specific conditional probabilities for the five-class solution of all MTA.

## Appendix C Alternative Latent Class Solutions

This appendix presents the four-class and six-class latent class analysis solutions that were considered in addition to the primary five-class model presented in the main text.

**Table A3:** Class specific conditional probabilities for the four-class solution of MTA patients with prehospital physiology.

| **Variable** | **Response** | **Class 1** | **Class 2** | **Class 3** | **Class 4** |
| --- | --- | --- | --- | --- | --- |
| Assigned Class Counts | N patients | 609 | 1221 | 908 | 1665 |
| Model Estimated Probabilities | NA | 0.144 | 0.276 | 0.204 | 0.376 |
| **ISS** | $ISS \leq 8$ | 0.079 | 0.192 | 0.012 | 0.313 |
| | $9 \leq ISS \leq 15$ | 0.142 | 0.458 | 0.988 | 0.334 |
| | $ISS > 15$ | 0.779 | 0.350 | 0.000 | 0.353 |
| **Age (years)** | $\leq 35$ | 0.203 | 0.251 | 0.020 | 0.001 |
|  | 36-55 | 0.271 | 0.374 | 0.145 | 0.014 |
|  | 56-65 | 0.150 | 0.183 | 0.349 | 0.071 |
|  | 66-80 | 0.279 | 0.172 | 0.195 | 0.353 |
| | $> 80$ | 0.098 | 0.020 | 0.291 | 0.561 |
| **Sex** | Female | 0.278 | 0.213 | 0.664 | 0.609 |
|  | Male | 0.722 | 0.787 | 0.336 | 0.391 |
| **Most Senior**<br>**Person**<br>**in the ED** | Consultant | 0.447 | 0.231 | 0.104 | 0.162 |
|  | Registrar | 0.315 | 0.464 | 0.526 | 0.536 |
|  | Senior House Officer | 0.038 | 0.060 | 0.191 | 0.150 |
|  | Specialist Registrar | 0.200 | 0.246 | 0.179 | 0.152 |
| **Most Severely**<br>**Injured**<br>**Body Area** | Chest | 0.091 | 0.291 | 0.000 | 0.178 |
|  | Head | 0.425 | 0.177 | 0.000 | 0.291 |
|  | Limbs | 0.100 | 0.183 | 0.993 | 0.168 |
|  | Spine | 0.160 | 0.137 | 0.007 | 0.249 |
|  | Other | 0.225 | 0.212 | 0.000 | 0.114 |
| **Transfers** | 0 | 0.343 | 0.959 | 0.969 | 0.976 |
|  | 1 | 0.109 | 0.000 | 0.001 | 0.008 |
|  | 2+ | 0.548 | 0.041 | 0.030 | 0.016 |
| **Length of Stay**<br>**(days)** | Q1 ($\leq 5$) | 0.176 | 0.408 | 0.187 | 0.159 |
|  | Q2 (6-9) | 0.092 | 0.352 | 0.252 | 0.198 |
|  | Q3 (10-18) | 0.205 | 0.190 | 0.330 | 0.293 |
|  | Q4 (18+) | 0.527 | 0.051 | 0.231 | 0.349 |
| **Mechanism**<br>**of Injury** | Fall less than 2m | 0.329 | 0.225 | 0.927 | 0.922 |
|  | Fall more than 2m | 0.220 | 0.226 | 0.020 | 0.044 |
|  | Other | 0.152 | 0.172 | 0.013 | 0.006 |
|  | Vehicle Incident | 0.298 | 0.378 | 0.041 | 0.028 |
| **Discharge**<br>**Destination** | Home based care | 0.214 | 0.957 | 0.536 | 0.469 |
|  | Institutional care | 0.039 | 0.026 | 0.270 | 0.335 |
|  | Mortuary | 0.593 | 0.002 | 0.164 | 0.099 |
|  | Rehabilitation | 0.154 | 0.014 | 0.030 | 0.097 |
| **Admitted to**<br>**Critical Care** | Yes | 0.697 | 0.102 | 0.021 | 0.024 |
|  | No | 0.303 | 0.898 | 0.979 | 0.976 |
| **Number of**<br>**Operations** | 0 | 0.371 | 0.674 | 0.000 | 0.939 |
|  | 1 | 0.337 | 0.201 | 0.809 | 0.049 |
|  | 2+ | 0.292 | 0.125 | 0.191 | 0.012 |

**Table A4:** Class specific conditional probabilities for the six-class solution of MTA patients with prehospital physiology.

| Variable | Response | Class 1 | Class 2 | Class 3 | Class 4 | Class 5 | Class 6 |
| --- | --- | --- | --- | --- | --- | --- | --- |
| Assigned Class Counts | N patients | 607 | 863 | 510 | 869 | 416 | 1138 |
| Model Estimated Probabilities | NA | 0.143 | 0.197 | 0.119 | 0.193 | 0.094 | 0.254 |
| **ISS** | ISS ≤ 8 | 0.003 | 0.271 | 0.020 | 0.008 | 0.103 | 0.462 |
| | 9 ≤ ISS ≤ 15 | 0.117 | 0.620 | 0.131 | 0.988 | 0.175 | 0.440 |
|  | ISS > 15 | 0.879 | 0.109 | 0.849 | 0.004 | 0.722 | 0.098 |
| **Age (years)** | ≤ 35 | 0.013 | 0.232 | 0.269 | 0.014 | 0.214 | 0.001 |
|  | 36-55 | 0.078 | 0.377 | 0.321 | 0.133 | 0.281 | 0.005 |
|  | 56-65 | 0.075 | 0.214 | 0.121 | 0.351 | 0.168 | 0.075 |
|  | 66-80 | 0.366 | 0.151 | 0.263 | 0.199 | 0.236 | 0.340 |
|  | > 80 | 0.469 | 0.025 | 0.026 | 0.303 | 0.101 | 0.579 |
| **Sex** | Female | 0.482 | 0.223 | 0.226 | 0.683 | 0.270 | 0.657 |
|  | Male | 0.518 | 0.777 | 0.774 | 0.317 | 0.730 | 0.343 |
| **Most Senior**<br>**Person**<br>**in the ED** | Consultant | 0.191 | 0.185 | 0.438 | 0.101 | 0.385 | 0.152 |
|  | Registrar | 0.542 | 0.501 | 0.278 | 0.532 | 0.384 | 0.528 |
|  | Senior House Officer | 0.112 | 0.080 | 0.010 | 0.195 | 0.053 | 0.164 |
|  | Specialist Registrar | 0.155 | 0.234 | 0.275 | 0.172 | 0.177 | 0.155 |
| **Most Severely**<br>**Injured**<br>**Body Area** | Chest | 0.062 | 0.293 | 0.237 | 0.000 | 0.071 | 0.230 |
|  | Head | 0.910 | 0.036 | 0.384 | 0.000 | 0.394 | 0.000 |
|  | Limbs | 0.023 | 0.300 | 0.020 | 0.998 | 0.138 | 0.235 |
|  | Spine | 0.000 | 0.203 | 0.009 | 0.002 | 0.228 | 0.367 |
|  | Other | 0.005 | 0.168 | 0.350 | 0.000 | 0.168 | 0.167 |
| **Transfers** | 0 | 0.989 | 0.950 | 0.932 | 0.972 | 0.069 | 0.966 |
|  | 1 | 0.009 | 0.001 | 0.006 | 0.001 | 0.149 | 0.010 |
|  | 2+ | 0.001 | 0.049 | 0.062 | 0.027 | 0.783 | 0.024 |
| **Length of Stay**<br>**(days)** | Q1 (≤ 5) | 0.268 | 0.458 | 0.324 | 0.170 | 0.088 | 0.110 |
|  | Q2 (6-9) | 0.199 | 0.355 | 0.234 | 0.249 | 0.096 | 0.208 |
|  | Q3 (10-18) | 0.243 | 0.157 | 0.254 | 0.340 | 0.200 | 0.312 |
|  | Q4 (18+) | 0.290 | 0.030 | 0.188 | 0.241 | 0.616 | 0.370 |
| **Mechanism**<br>**of Injury** | Fall less than 2m | 0.897 | 0.241 | 0.147 | 0.949 | 0.366 | 0.925 |
|  | Fall more than 2m | 0.081 | 0.221 | 0.235 | 0.014 | 0.212 | 0.034 |
|  | Other | 0.009 | 0.145 | 0.250 | 0.009 | 0.115 | 0.008 |
|  | Vehicle Incident | 0.014 | 0.393 | 0.368 | 0.028 | 0.307 | 0.033 |
| **Discharge**<br>**Destination** | Home based care | 0.524 | 0.963 | 0.747 | 0.518 | 0.117 | 0.457 |
|  | Institutional care | 0.239 | 0.030 | 0.060 | 0.281 | 0.005 | 0.363 |
|  | Mortuary | 0.063 | 0.007 | 0.021 | 0.170 | 0.849 | 0.122 |
|  | Rehabilitation | 0.174 | 0.000 | 0.173 | 0.032 | 0.029 | 0.058 |
| **Admitted to**<br>**Critical Care** | Yes | 0.050 | 0.022 | 0.473 | 0.023 | 0.673 | 0.024 |
|  | No | 0.950 | 0.978 | 0.527 | 0.977 | 0.327 | 0.976 |
| **Number of**<br>**Operations** | 0 | 0.946 | 0.601 | 0.698 | 0.000 | 0.212 | 0.931 |
|  | 1 | 0.035 | 0.257 | 0.158 | 0.811 | 0.439 | 0.058 |
|  | 2+ | 0.019 | 0.142 | 0.144 | 0.189 | 0.349 | 0.011 |

